# Establishment of a large-scale oral disease registry (NDCS-ODR) in a national specialty center

**DOI:** 10.64898/2026.01.13.26344086

**Authors:** John Rong Hao Tay, Gustavo G Nascimento, Jasmine Sio Hong Ho, Narayanan Ragavendran, Bryan Wei Ren Yeo, Hanimi Reddy Kallam, Sean Shao Wei Lam, Marco A Peres

## Abstract

This study describes the establishment of the National Dental Centre Singapore Oral Disease Registry (NDCS-ODR), a large-scale, electronic health records registry designed to capture real-world data on oral diseases. The NDCS-ODR was developed to standardize and integrate oral health data within Singapore Health Services, the country’s largest healthcare cluster. Its development, governance, and data architecture are described, with an overview of individuals with oral diseases recorded in the registry. Data collection from 2013 to June 2025 has been completed. As of June 2025, the NDCS-ODR comprises 229,249 unique patients, with a mean (SD) age of 49.1 (19.5) years and an approximately equal sex distribution. Most were of Chinese ethnicity (77.6%), and Singapore citizens (92.5%). Clinical variables indicated substantial disease and treatment burden, with a mean of 7.9 (7.7) missing teeth, 4.8 (6.2) restored surfaces, and 2.8 (3.4) restored teeth per patient. Among 108,517 recorded periodontal diagnoses, Stage III periodontitis (2018 EFP/AAP Classification) and severe chronic periodontitis (1999 Classification of Periodontal Diseases and Conditions) were most common. The NDCS-ODR represents Singapore’s first large-scale, real-world oral disease registry embedded within a national specialty center, demonstrating the feasibility of leveraging electronic health record data for research and service evaluation.

## Introduction

Oral diseases are among the most prevalent non-communicable diseases worldwide [1]. The World Health Organization estimates that nearly 3.7 billion people, half of the world population, are affected by untreated caries, periodontitis, edentulism, and other oral disorders. Furthermore, caries and periodontitis are the two most common oral diseases and major causes of tooth loss, and can compromise nutrition and reduce quality of life [2, 3]. Their consequences extend beyond physical health, influencing psychosocial well-being and increasing mortality [4-9]. By 2050, the global burden is projected to rise substantially, with severe periodontitis affecting more than 1.5 billion people and edentulism exceeding 660 million cases [10]. Similarly, the prevalence of permanent caries is projected to increase by more than 20% [11]. Poor oral health also exerts a considerable economic burden worldwide; in 2019, the combined global costs of oral diseases were estimated at approximately US$710 billion, reflecting substantial healthcare spending on treatment as well as productivity losses [12].

In Singapore, oral health challenges remain significant. Although complete edentulism affects about 13% of adults aged 60 years and above, a recent national survey reported that dental caries affects 85% of the population, with 35% having at least one untreated carious lesion [13, 14]. Periodontitis is highly prevalent, affecting 78% of the population, with 16% presenting with its severe form [13]. As one of the fastest-ageing societies globally [15], preventing tooth loss in its population is a key factor for preserving nutrition, well-being, and healthy longevity.

High-quality real-world data are essential for understanding the determinants and outcomes of oral disease, evaluating the quality of care, informing healthcare policy, and planning the future workforce. In oral health, routinely collected clinical data from electronic health records provide data that enable benchmarking of outcomes, detection of unwarranted variation, and the targeting of service improvements toward populations and practices with the greatest need [16-18]. Singapore Health Services (SingHealth) functions as Singapore’s largest healthcare cluster and operates as an Academic Medical Centre. Within this network, initiatives such as the Perioperative and Anesthesia Subject Area Registry (PASAR) [19], the SingHealth Diabetes Registry (SDR) [20], and the SingHealth COPD and Asthma Data Mart (SCDM) [21] illustrate how electronic health record–integrated registries can advance observational research, quality improvement, and data-driven innovation. These systems rely on data extraction from electronic health records, harmonized data architectures, rigorous quality assurance, and privacy-preserving de-identification processes. Establishing a comparable infrastructure for oral health would allow systematic study of disease patterns, trajectories, and clinical outcomes in routine care settings.

SingHealth’s enterprise information technology ecosystem already supports a wide spectrum of administrative, clinical, and operational needs through its data warehouse, known as the Electronic Health Intelligence System (eHints), which consolidates data across SingHealth institutions [22]. Leveraging this framework would facilitate the linkage and integration of oral health data with other health records, enabling the investigation of oral and broader health outcomes.

The National Dental Centre Singapore (NDCS) is the country’s flagship specialist center for oral healthcare. It is the largest oral healthcare center in Singapore and one of five national specialty centers within the SingHealth cluster. By utilizing NDCS’s electronic health records, the NDCS Oral Disease Registry (NDCS-ODR; hereafter referred to as ODR) catalogs diagnoses, procedures, and outcomes for individuals receiving care at the center. This paper introduces the methodology for establishing the ODR. The objectives are: (1) to provide an overview of individuals with oral diseases captured in the ODR, (2) to describe the technical and governance framework used to create the real-world data repository, and (3) to illustrate its application through a focused evaluation of data accuracy and completeness for periodontal records between 2013 and 2025. Periodontal disease was selected as the initial focus because it combines structured data with considerable analytical complexity. Accurate periodontal surveillance relies on tooth-level measurements recorded at multiple sites per tooth, which are often incomplete, require examiner calibration to minimize measurement variability, and must account for a major shift in diagnostic frameworks over the observation period. Its strong relevance to tooth survival and systemic health outcomes further makes it a clinically valuable first test of the ODR’s capacity to support epidemiology, quality evaluation, and cross-registry linkage.

## Materials and Methods

### Source system

The registry’s data source is the proprietary Electronic Dental Record (EDR) used at NDCS—developed by Integrated Health Information Systems (IHiS, now known as Synapxe). The extracted registry’s domain-specific EDR data captures structured clinical information, including demographics, diagnoses, procedures, tooth condition, and periodontal charting (probing pocket depth [PPD], recession [REC] and bleeding on probing [BOP]). It does not include radiographic, administrative or pharmacy data, and free text notes are not extracted for the registry. Figure 1 illustrates the data architecture and workflow from EDR to ODR.

**Fig 1.** Data architecture. **Abbreviations:** DB, database; EDR, Electronic Dental Record; ODR, Oral Disease Registry.

### Administration and governance

The ODR currently includes all patients who had a dental visit at NDCS between 01/01/2013 and 30/06/2025. The registry was established under CIRB exemption 2022/2167 with a waiver of informed consent because only routinely collected, anonymized clinical data were used. Governance follows Singapore’s Personal Data Protection Act, the Human Biomedical Research Act (HBRA), and the Declaration of Helsinki. Each patient is assigned a unique identifier (NRIC for citizens and permanent residents), which serves as a linkage key across visits and enables integration with other healthcare databases within SingHealth. NRICs were encrypted before storage; a linkage file permitting re-identification requires approval from the SingHealth Office of Deputy Group Chief Medical Informatics Officer (Research); and all data are stored in secure, access-controlled environments. Analysts access de-identified data through virtual desktops within the SingHealth network using corporate credentials and devices, ensuring that no data are stored or processed outside the secure infrastructure.

A formal governance process has been established for the access, storage, and use of the data. All approved data are stored within the SingHealth Research Control Tower, a secure, cloud-based institutional data platform. Access is restricted to the institutional network via a secure platform, and the system is commercially hosted in compliance with institutional data governance and security policies. Access is limited to authorized users through role-based permissions, and storage of data on personal devices is not permitted unless explicitly approved and secured in accordance with institutional guidelines. Researchers seeking to use ODR data follow a structured and transparent process. They begin by submitting a proposal outlining their intended analysis, which is shared with ODR investigators for feedback and potential collaboration. Once endorsed, the researcher submits a formal data request to the ODR Executive Committee for approval. External collaborators may also participate by contacting the ODR Principal Investigator, who reviews and circulates the proposal among ODR investigators for consideration.

### System development

Initial discussions were held with the Principal Investigator, data manager, NDCS Chief Medical Informatics Officer, and Synapxe engineers to define the project scope and data requirements. EDR Subject area variables were identified and mapped to EDR’s metadata repository or data dictionary to understand the relationships between its elements to other data, usage and formats. Screens from the NDCS EDR were systematically reviewed to identify and confirm the exact front-end fields required for extraction at record-level from a larger dataset. These were subsequently traced to their corresponding back-end tables and compiled into a User Requirements Document outlining clinical variables and inclusion rules, and a Functional and Technical Specification detailing data elements, linkage keys (tooth, site, and patient level), and data flow architecture.

The team designed the registry schema and implemented a two-step extract–transform–load process:

(i) Cohort identification, which defined eligibility criteria (i.e., patient with an initial assessment visit note), and
(ii) Data import and mapping, which integrated the relevant clinical elements.

A data harmonization step reconciled discrepancies between front-end variable names and back-end storage formats. ETL scripts were implemented and validated through internal integration testing to ensure data lineage and field-level accuracy. User acceptance testing involved three independent testers—two clinicians and a data manager—who evaluated both functionality and data fidelity using a synthetic test cohort of 45 mock patients, each with 10 scheduled visits. The UAT primarily focuses on validating data constraints and rules established across the EDR datasets, checking on the consistency and completeness of data to ensure data integrity is maintained. Test visits were distributed across fixed weekdays to emulate real operational patterns such as clinic scheduling and end-of-day data loads. Testing comprised three key domains: a) functional verification: ensuring screens and queries retrieved the expected fields, and that diagnostic and odontogram logic operated as designed (the odontogram refers to the graphical charting interface in the EDR that records each tooth’s surface condition and periodontal charting; b) data verification: confirming that record counts and patient-timepoints in the ODR matched EDR extracts under identical inclusion criteria, with field-level checks on presence, surface condition, PPD, REC, BOP, and c) soak testing: performed 10 weeks after UAT initiation to confirm that newly generated visits on the scheduled days were correctly captured by the ETL runs, simulating ongoing production operations.

### Data collected

For each eligible visit, the ODR captured:

- Demographics: age, date of birth, sex, ethnicity, residency status, Singapore Housing Index, a proxy measure of socioeconomic position based on the type of public housing (e.g., 1-2 room, 3 room, 4-5 room, or private housing) [23].
- Smoking history
- Diagnosis codes
- Examination chart variables: complaints, dental history, extra-oral and intra-oral findings
- Odontogram data: presence or absence of each tooth, tooth and surface condition (including caries, restorations, crowns, root-filled teeth, and prosthetic status), and periodontal charting (PD, REC and BOP for six sites per tooth)

### Data quality assessment

#### Inclusion criteria and extraction rules for periodontal cases

Records entered the registry for periodontal cases through the ETL process if they had: (1) an initial assessment visit or periodontics treatment note; (2) a periodontal diagnosis recorded; (3) non-empty odontogram data; and (4) odontogram data that match the visit date within ±1□day. Variables extracted included tooth presence/absence, surface conditions, PPD, REC, BOP, treatment codes, demographics, and smoking status.

#### Accuracy of periodontitis classification

To assess diagnostic accuracy of periodontitis cases, the registry randomly sampled records meeting two criteria: (1) complete periodontal charting (PD, REC and BOP) and (2) a recorded periodontal diagnosis from patients with a documented periodontal treatment note, indicating clinical management within the periodontal unit at NDCS. Each selected case was independently classified by a calibrated examiner using only the periodontal charting data, blinded to the original recorded diagnosis. Classification criteria were clarified in advance through discussions with a senior epidemiologist to ensure consistency. Agreement between the examiner’s classification and the registry diagnosis was then assessed.

Two diagnostic systems were used over the observation period: the 1999 Classification of Periodontal Diseases and Conditions (CPDC) classification (used up to April 2019), and the 2018 European Federation of Periodontology/American Academy of Periodontology (EFP/AAP) classification (used from May 2019). For the latter, only staging was assessed due to a lack of radiographs. Classifications for 1999 CPDC follow Armitage [24], while those for the 2018 EFP/AAP classification follow Tonetti et al [25, 26]. The electronic dental records in NDCS use a structured diagnostic field with predefined dropdown options, which were updated from the 1999 CPDC to the 2018 EFP/AAP classification in May 2019. Prior to this transition, discussions on the application of the new staging and grading framework were conducted within the periodontal unit, and the EFP clinical decision tree for periodontitis staging and grading [27] was disseminated to all clinicians at the center as a standardized diagnostic reference. Formal inter-examiner calibration exercises for diagnostic coding were not conducted; instead, diagnostic standardization relied on the structured data entry system, the clinical decision tree, and consultant oversight within the periodontal unit. Notably, agreement rates were similar across periods using the 1999 CPDC and the 2018 EFP/AAP classifications (Table 4), suggesting that the transition between classification systems did not substantively affect diagnostic reliability within the registry. Power calculations indicated that 35 cases per period were sufficient to estimate a 90% agreement rate with 95% confidence. Consequently, 35 cases were randomly sampled within each period, resulting in a total of 140 cases stratified approximately equally across four periods to account for clinician turnover and potential variations in diagnostic practices: 01/01/2013-29/02/2016 (38 months), 31/03/2016-30/04/2019 (38 months), 01/05/2019-30/06/2022 (38 months), and 31/07//2022-30/06/2025 (36 months).

#### Completeness of periodontal charting

Completeness was assessed at both the site– and patient-level. The analysis began with quantifying the total number of missing sites across the entire dataset to provide an overall measure of data quality. The proportion of records with 100% charting completeness was then determined, where completeness was defined as non-missing values for probing depth (PD), gingival recession (REC), and bleeding on probing (BOP) at all six sites per present tooth. To examine changes in charting quality over the course of treatment per patient, the percentage of patients with fully complete charts at their first two visits was evaluated. Analyses for the current phase are limited to the first two visits per patient. Future updates will incorporate additional visits once further data extraction is completed. In parallel, a clustered logistic factor multiple-imputation approach was developed and validated to address incomplete periodontal charting in this registry[28]. A masking experiment was conducted on complete charts: sites were artificially removed and then imputed to evaluate performance against the known truth, and imputation of binary disease indicators (e.g., probing depth ≥5 mm, clinical attachment level ≥3 mm) was done by modeling within-examination spatial dependence, enabling bias-reduced estimation of disease prevalence and longitudinal patterns from routinely collected data. For the purposes of this study, the NDCS-ODR dataset used for this study was extracted and accessed for analysis on 04/04/2025 and last accessed on 15/12/2025. The study team accessed de-identified registry data only; directly identifying information was not accessible to the study team during or after data extraction/analysis.

## Results

### Characteristics of the ODR cohort

From 2013 to mid-2025, the ODR captured 229,249 unique patients (Table 1). The sex distribution was roughly balanced throughout, and the cohort’s composition was predominantly Chinese (∼78%) and Singapore citizens (∼93%). The cohort aged over time: mean age rose steadily from the late-40s in the to the mid-50s by 2023–2025. Smoking status was dominated by never-smokers across all years, with a modest decline in current smoking over time. Most clinical variables showed right-skewed distributions: median values were zero and means were low, indicating that while most patients had no recorded events for these measures, a smaller proportion exhibited higher counts (Table 2).

**Table 1.**
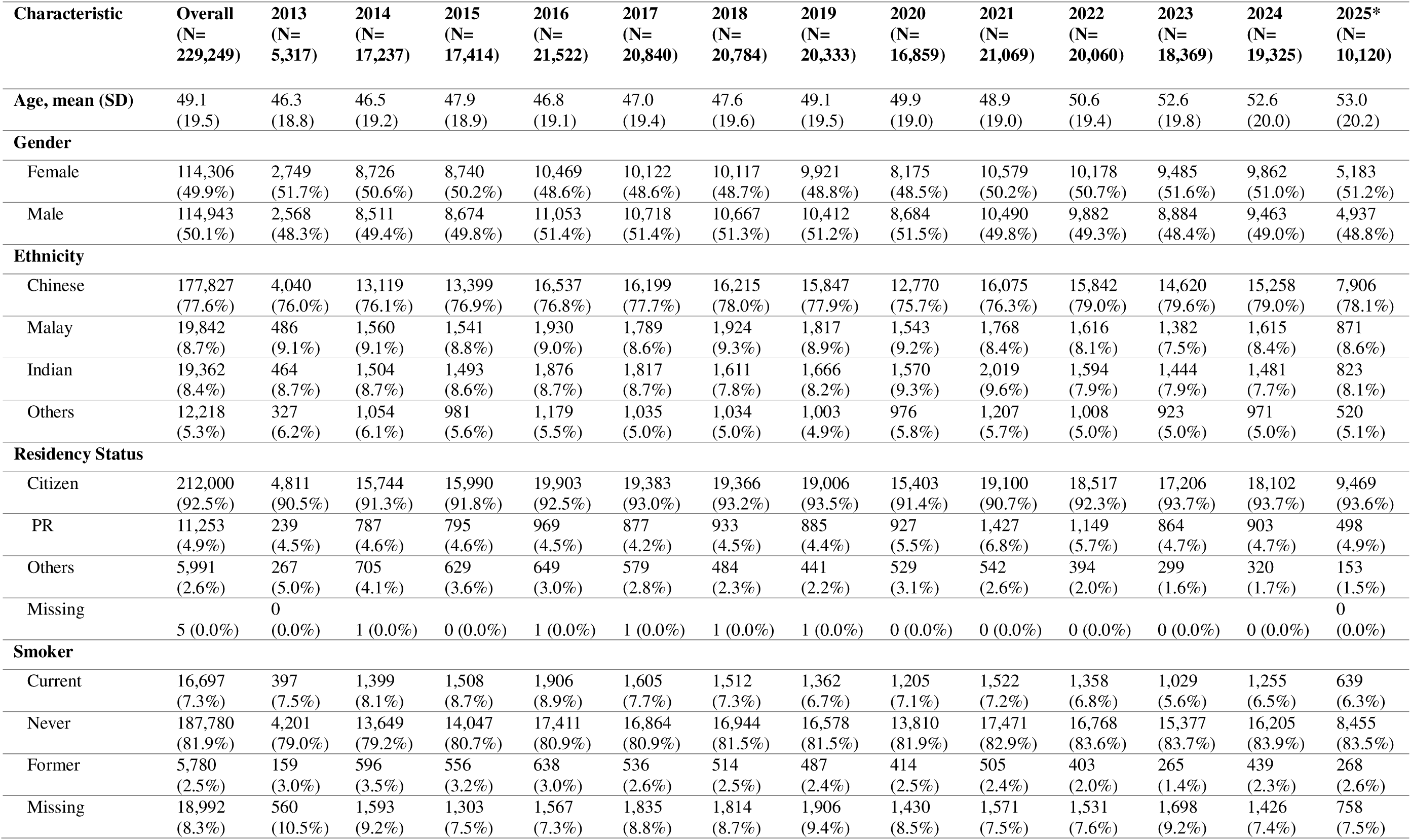
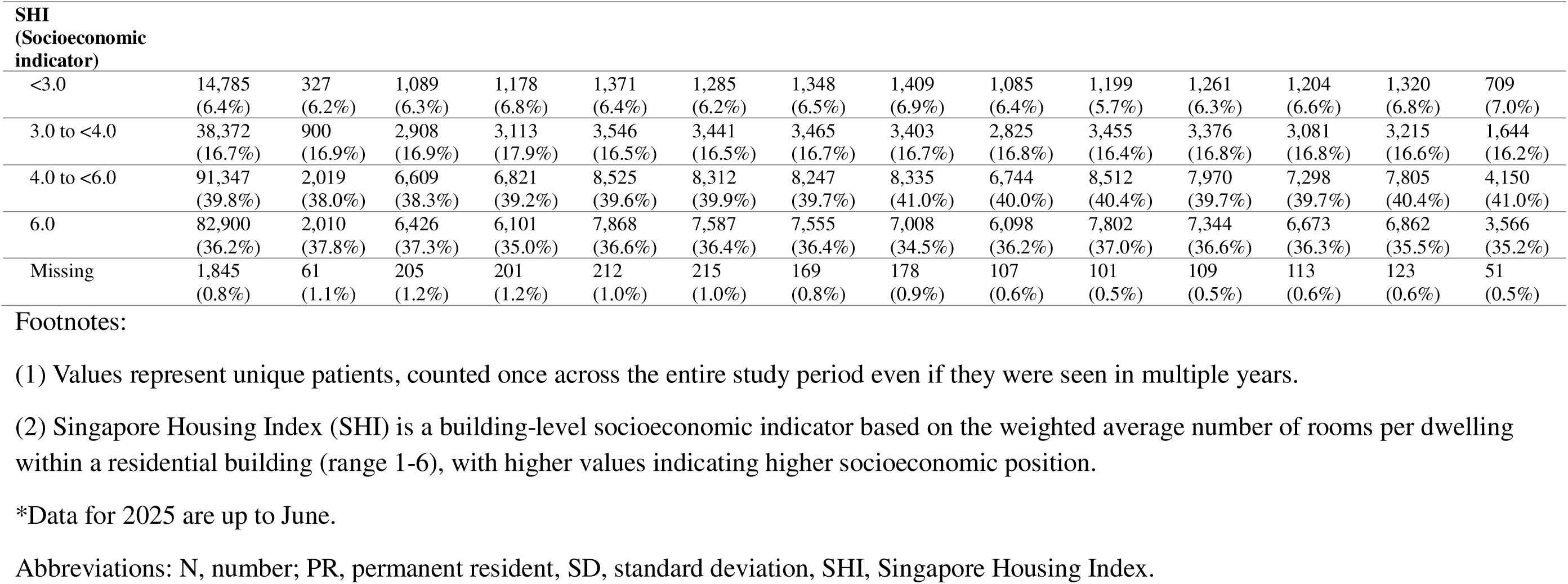
Characteristics of patients in the NDCS Oral Disease Registry, 2013–2025.

**Table 2.**
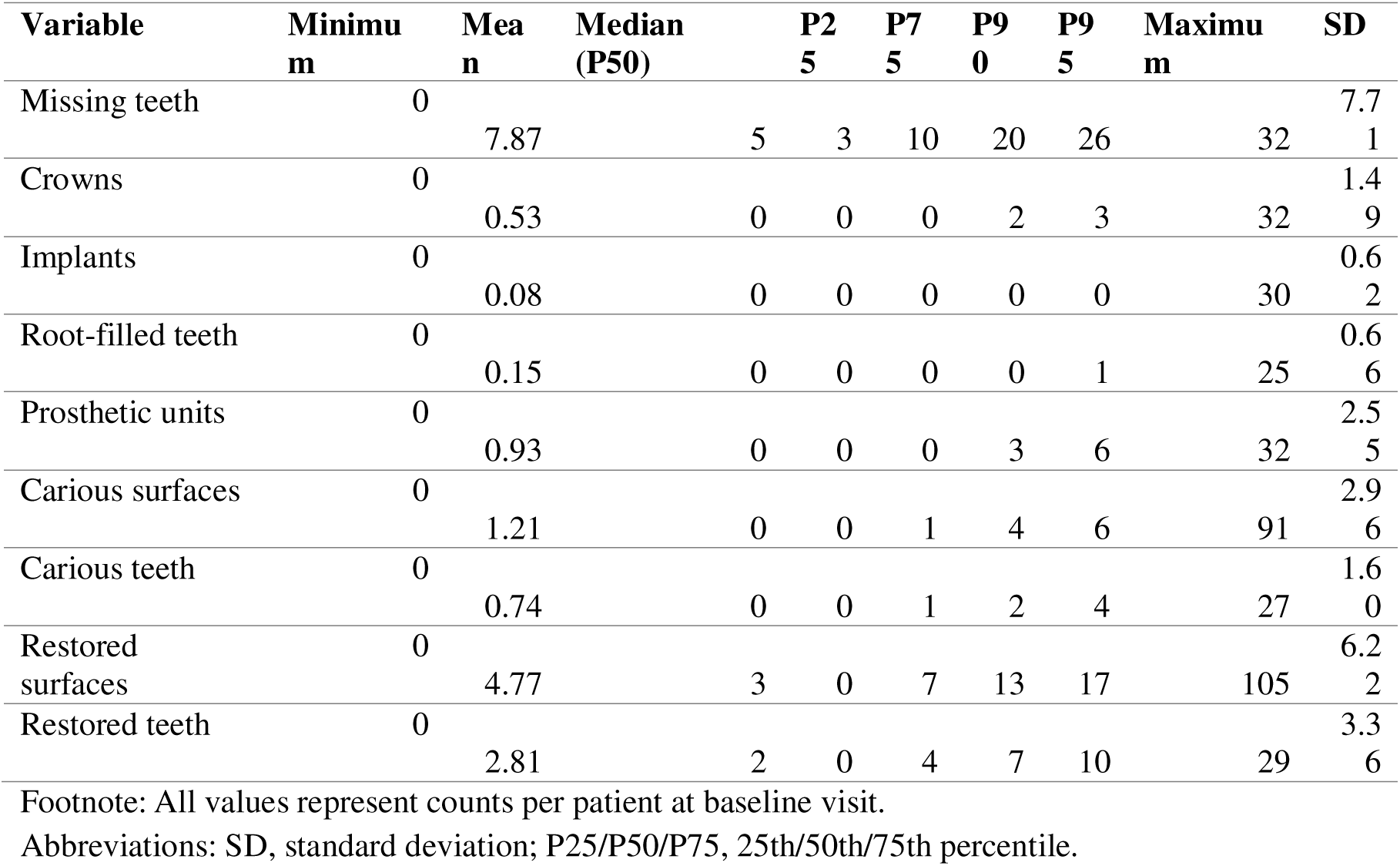
Summary statistics of clinical variables recorded.

### Periodontal diagnoses and service activity

The overall distribution of periodontal diagnoses is presented in Table 3, with the full list in S1 Table. Among cases classified with the 2018 EFP/AAP system, Stage III disease predominated, with both localized and generalized presentations represented across grades. For diagnoses recorded under the 1999 CPDC Classification, localized and generalized severe chronic periodontitis were the most common categories. Periodontal service activity by year and visit type is summarized in S2 Table. Visit counts increased over time, with relatively lower counts observed in the earlier years of the registry period, followed by a marked increase from 2019 onwards. A transient drop in visits was observed in 2020, consistent with reduced clinical activity during the COVID-19 pandemic, followed by recovery in subsequent years.

**Table 3.** Distribution of periodontal diagnoses in the ODR.

| Diagnosis category | N | % |
| --- | --- | --- |
| EFP/AAP Stage I–II periodontitis | 2,814 | 2.59 |
| EFP/AAP Stage III periodontitis | 27,051 | 24.93 |
| EFP/AAP Stage IV periodontitis | 9,188 | 8.47 |
| CPDC severe chronic periodontitis | 13,131 | 12.10 |
| CPDC mild/moderate chronic periodontitis | 8,542 | 7.87 |
| CPDC aggressive periodontitis | 460 | 0.42 |
| Gingivitis (localized/generalized) | 9,995 | 9.21 |
| Peri-implantitis/Peri-implant mucositis | 2970 | 2.74 |
| Acute periodontal lesions (abscess / endo-periodontal) | 5,896 | 5.43 |
| Other periodontal conditions* | 5,603 | 5.16 |
| Unspecified periodontal conditions | 22867 | 21.07 |
| **Overall** | **108,517** | **100.00** |

Footnote: All values represent counts per patient at baseline visit. Other periodontal conditions include drug-induced gingival overgrowth; occlusal trauma; gingival recession; periodontitis modified by systemic disease. See S1 Table for a full listing and counts.
Abbreviations: CPDC, 1999 Classification of Periodontal Diseases and Conditions classification; EFP/AAP, 2018 European Federation of Periodontology/American Academy of Periodontology classification.

### Diagnosis agreement

Agreement between examiner-derived and recorded periodontal diagnoses varied by period (Table 4). The highest concordance was observed during the final years when the 1999 CPDC classification was still in use and in the most recent period, with lower agreement noted in the earliest ODR years and during the initial implementation of the 2018 EFP/AAP classification. After combining all diagnoses, unweighted Cohen’s kappa indicated substantial agreement: κ = 0.84 (95% CI 0.78–0.90).

**Table 4.** Agreement between examiner-derived and recorded periodontal diagnoses across three time periods.

| Time Period | Classification System | N | Agreement (%) | 95% CI |
| --- | --- | --- | --- | --- |
| January 2013–February 2016 | 1999 CPDC | 35 | 80.0 | 66.7–93.3 |
| March 2016–April 2019 | 1999 CPDC | 35 | 91.4 | 82.2–100.0 |
| May 2019–June 2022 | 2018 EFP/AAP | 35 | 82.9 | 70.4–95.3 |
| July 2022–June 2025 | 2018 EFP/AAP | 35 | 91.4 | 82.2–100.0 |

Abbreviations: CI, confidence interval; CPDC, 1999 Classification of Periodontal Diseases and Conditions classification; EFP/AAP, 2018 European Federation of Periodontology/American Academy of Periodontology classification; N, number.

### Data completeness

Data completeness was low, with REC showing the highest proportion of missing data (Table 5). Completeness improved at follow-up compared with initial visits, likely because second visits typically corresponded to periodontal treatment appointments, who recorded more comprehensive charting. Nevertheless, PPD and BOP were only slightly more than half complete. At the site level, REC similarly exhibited the greatest degree of incompleteness across parameters (S3 Table).

**Table 5.**
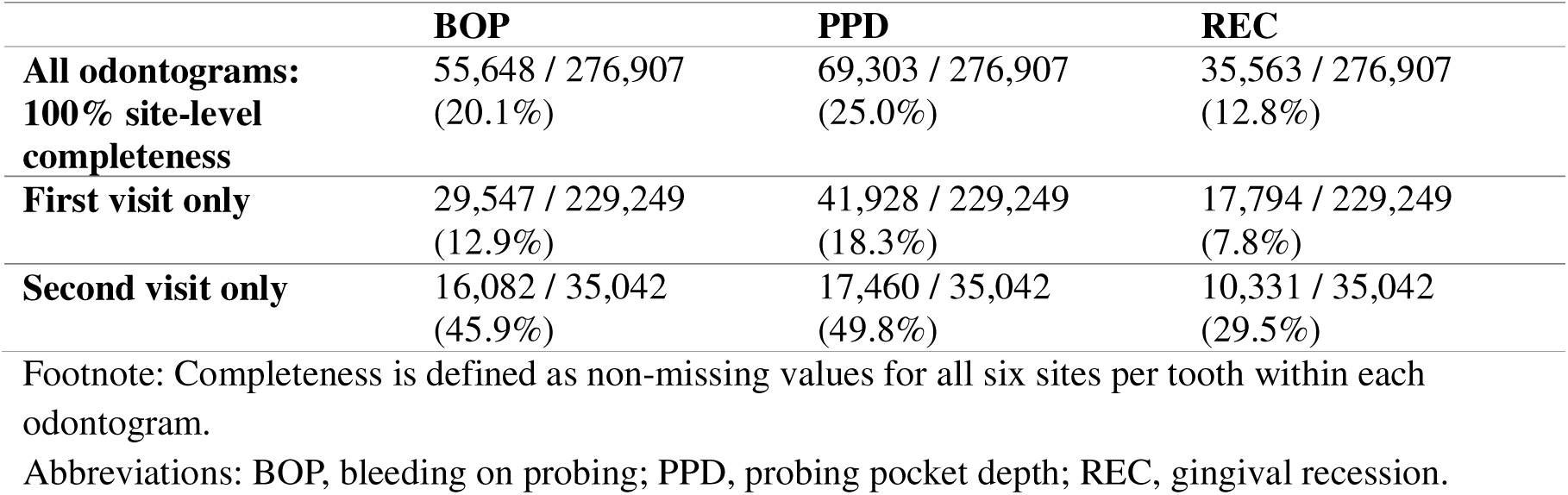
Completeness of periodontal charting.

## Discussion

This study describes the establishment of the ODR, including its governance, data architecture, and the processes used to extract and harmonize structured electronic dental record data between 2013 and 2025. In the focused periodontal evaluation, examiner-derived and recorded diagnoses showed substantial agreement, supporting the use of registry-coded periodontitis diagnoses for epidemiologic analyses. However, periodontal charting completeness—particularly for gingival recession—was limited, indicating that analyses relying on full clinical charting may be affected by under-recording and should incorporate strategies to address missing data and documentation variability.

Routine electronic health records accumulate large volumes of patient data and can be utilized to generate real□world evidence about oral diseases. There is increasing recognition that secondary use of electronic health record data can improve the knowledge base in dentistry, enabling questions to be addressed regarding treatment effectiveness and the links between oral and overall health [29, 30]. Clinical trials, while rigorous, are costly and often involve highly selective populations that may not reflect real-world treatment effects [31]. Administrative or claims databases, on the other hand, capture all patient interactions with the healthcare system but lack important clinical details such as examination findings, diagnostic test results, and prescribed medications [32]. Administrative databases are also limited in their ability to explore clinical and contextual factors underlying oral health disparities, such as differences in disease burden and access to care across population groups [33]. Although data leakage or misclassification can occur, electronic health records occupy a middle ground by combining the clinical richness of chart data with the large scale that claims data provide.

Although large-scale electronic dental record repositories are still relatively uncommon globally, several initiatives show that routinely collected dental data can be curated for epidemiological and outcomes research. In the United States, the BigMouth Dental Data Repository was established in 2012 using electronic data from four academic dental schools; it initially aggregated 1.1 million patient records[29], and has since expanded to 18 schools with over 6 million records [34]. Practice-based research networks have also demonstrated the feasibility of using standardised electronic dental record data across distributed clinics for multi-site observational studies [35]. In Nordic countries, national dental registries—such as Denmark’s National Child Odontology Registry [36] and Sweden’s Dental Health Register [37]—further illustrate how structured dental data can support population surveillance research.

In contrast, the ODR is a single□center registry based on structured data. It leverages SingHealth’s enterprise data infrastructure for warehousing and governance, but currently encompasses only one of Singapore’s two national specialty centers. Despite its narrower scope, the ODR includes more than 229,000 unique patients over 12 years and provides detailed, tooth– and site-level odontogram data on caries and periodontal status. As the ODR is embedded within an integrated health system, it enables linkage of oral health data with medical databases, capabilities not available in existing dental repositories such as BigMouth. Therefore, the ODR provides unique opportunities to study oral and systemic relationships.

The ODR has approximately equal numbers of male and female patients and an ethnic distribution comprising predominantly Chinese individuals. The mean age increased over the study period, suggesting that older adults are increasingly seeking specialist oral care. These demographic trends are broadly consistent with findings from national-representative oral health surveys [13], although the ODR included fewer Malay and permanent-resident patients, and proportionally more citizens. The differences in residency status compared to Singapore proportion are likely attributable to referral pathways and subsidy structures within the public healthcare system. Clinical variables were markedly skewed, with median counts for many parameters at zero and means close to zero. This likely reflects a mix of patients with low prior treatment burden and those attending for initial assessment or treatment planning, where treatment needs had not yet been recorded. However, the ODR should not be interpreted as population-representative of Singapore. As a specialist referral center, the registry captures individuals with generally higher disease burden and treatment complexity than would be observed in primary care or community settings, as well as those who can afford and are willing to seek specialist treatment. Conditions more likely managed in the primary care sector, such as early-stage caries or Stage I/II periodontitis, are likely underrepresented. Conversely, the registry provides a detailed characterization of moderate-to-severe disease that is not typically available from national surveys, which rely on partial-mouth examination protocols [13]. The ODR and population-based surveys [38] should therefore be viewed as complementary data sources, each addressing different segments of the disease spectrum.

Only about one□quarter of odontograms had complete charting for PPD and one□fifth for bleeding on probing (BOP), and just 12.8% were complete for REC. These findings are broadly consistent with evaluations from other dental repositories such as BigMouth, which similarly reported high completeness for demographic variables but variable completeness for clinical measures, often influenced by provider-level differences. Incomplete periodontal charting is associated with systematic underestimation of disease burden by 11-76% depending on the protocol and disease severity, with bias generally increasing for more severe case definitions [39, 40]. However, analyses using ODR periodontal records showed that multiple imputation can substantially reduce this bias, particularly for mild-to-moderate disease categories, while preserving overall diagnostic patterns observed in the registry [28]. Incomplete charting underestimated patient-level disease counts by approximately 13 sites per patient for clinical attachment level ≥3 mm, with smaller differences for deeper probing depth thresholds. Imputation effects were largest in less severe periodontitis categories, where site-level prevalence increased by 7-11 percentage points after imputation, compared with 3-5 percentage points for Stage III and IV periodontitis. These findings indicate that the primary bias from incomplete charting is systematic underestimation of mild-to-moderate disease burden, while estimates of advanced disease are relatively robust to missing data. These results should be interpreted with the caveat that multiple imputation assumes data are missing at random conditional on observed covariates. Sensitivity analyses under outcome-dependent missingness mechanisms showed that imputation remained substantially better than complete-case analysis, though residual bias persisted at the mildest disease thresholds. Agreement between examiner-derived and recorded diagnoses varied across time periods, with higher concordance observed in 2016–2019 and the most recent period. This likely reflected clinicians’ familiarity with the 1999 CPDC classification and ongoing calibration exercises linked to concurrent clinical studies. The subsequent dip in agreement may be explained by the transition to the newer 2018 EFP/AAP diagnostic framework and the associated learning curve during its early implementation. However, the overall level of agreement was substantial, demonstrating that periodontal diagnoses within the registry were largely concordant. This agreement estimate should be interpreted with the caveat that the validation sample was restricted to records with complete periodontal charting and a documented periodontal treatment note, which selects for patients who tolerated full six-point charting and, therefore, are more likely to be engaged in periodontal care. Diagnostic accuracy for cases with incomplete charting could not be assessed. These cases may reflect patients with lower treatment tolerance or more complex presentations, and agreement in such records may be lower.

### Lessons learnt

Electronic health records often operate in fragmented and inefficient systems, with limited interoperability and substantial data processing required to extract usable information. The lack of consistent structure and standardization further compounds these inefficiencies that hinder integration and secondary analysis [41]. Our experience revealed several operational challenges. First, initial assessment visit and treatment notes must be standardized and completed consistently. The relatively low number of recorded periodontal visits prior to 2019 likely reflects inconsistent use of structured periodontal note templates rather than true service volume. Second, data extraction depended on structured fields; free-text notes and radiographs were excluded, limiting capture of clinical data. Radiographs and prescriptions are stored in separate systems, further complicating data linkage.

Third, electronic health records are primarily designed for patient care rather than research, and substantial effort was required to harmonize variable names, map relationships from tooth to site to patient, and resolve inconsistencies. Finally, registry development requires close involvement of diverse stakeholders. This includes researchers, data managers, engineers, and clinicians familiar with the electronic health records system. Strong administrative leadership is also critical to navigate governance processes and ensure timely approvals. Clear governance structures and sufficient resource support are also necessary to maintain the registry’s long-term viability. The results from the registry can also be utilized for clinical audits, for example, by identifying gaps in documentation, and standardizing charting practices.

### Limitations and Strengths

The ODR is not without limitations. The ODR represents a single national specialty center and may not represent the broader community disease burden; the cohort is skewed toward citizens, and individuals seeking specialist care. Data completeness is limited. Whether missing charting data are outcome-dependent – for example, if clinicians are more likely to record sites with deeper pockets while omitting apparently healthy sites – cannot be determined from the registry, as the unrecorded values are by definition unknown. If such selective recording occurs, observed disease estimates may overestimate the true proportion across the full mouth and inflate the apparent prevalence of severe disease at the individual level. A particular challenge in dentistry is the inherently hierarchical structure of the oral cavity, where each patient has multiple teeth, each with several charted sites. This making comprehensive charting time-consuming in routine workflows. Moreover, the 1999 CPDC and 2018 EFP/AAP classifications require complete site-level data, a standard difficult to achieve in everyday clinical practice. Standardizing charting procedures and clinician training at NDCS will therefore be essential to improve data completeness and diagnostic reliability. A substantial proportion of visits carried unspecified periodontal codes, which is expected as many of these encounters were likely managed by clinicians from other specialties as part of routine triage, rather than by periodontists assigning definitive diagnoses. Smoking history was self-reported and may also have been under-recorded. Clinician-level identifiers are not captured in the ODR, precluding assessment of whether diagnostic coding differs across clinician groups. As a national specialty center operating within an academic medical center, cases managed within specialty units at NDCS, including the periodontal and restorative dentistry units, and are subject to consultant oversight, case presentations, and calibration discussions with general dental practitioners and residents, providing a degree of diagnostic standardization for both periodontal and caries diagnoses. However, encounters recorded during triage by clinicians from other dental specialties, which likely account for a substantial proportion of unspecified periodontal codes, do not undergo the same level of diagnostic review. Inter-clinician variability in recording practices may introduce information bias, particularly if diagnostic thresholds differ between specialists and generalists, for example, in the recording of early carious lesions or the assignment of periodontal case definitions.

Nevertheless, this study has strengths. It covers more than a decade of care for a large number of unique patients. The use of a unique national identifier for each citizen or permanent resident (NRIC) enables linkage to other SingHealth and national registries, providing opportunities to explore oral–systemic relationships. Standardization of data elements within the ODR enhances data quality and facilitates interoperability across healthcare domains. The calibrated diagnosis demonstrated substantial agreement, suggesting that clinical charting can be used reliably for epidemiological research.

Another strength of this study is the use of the SHI as a proxy for socioeconomic position [23]. In Singapore, home ownership and housing type serve as well-established socioeconomic indicators [42]. Individual-level data such as income or education are not routinely captured in electronic health records and may be subject to reporting and social desirability biases when self-reported. Area-level indices, such as neighborhood-based socioeconomic position measures, have been developed locally, but remain prone to ecological fallacy [43]. The SHI overcomes these limitations by providing building-level granularity, which retains privacy while approximating individual socio-economic status [44]. It is derived from open data Application Programming Interfaces (APIs) and represents a weighted average number of rooms per residential unit, scaled from 1 to 6. While it does not distinguish between private housing types (e.g., condominiums or landed properties), it preserves the ordinal housing-value gradient across the population (S4 Table). The SHI has been validated and shown to correlate with population-level indicators such as bystander cardiopulmonary resuscitation rates [45] and vaccination uptake [46].

### Future directions

To our knowledge, the ODR is the first large□scale oral disease registry in Asia established within a national specialty center. Through this, we hope to contribute to the growing literature on real□world oral health research and encourage collaborations that span registries and health systems. Ongoing work will extend the ODR to include data from other dental specialties (e.g., pediatric dentistry). Incorporation of clinical images and prescription data, as well as natural language processing of free□text notes will enhance data richness and research utility. We hope for the registry to be refreshed at six-month intervals, subject to operational feasibility and resource availability.

The ODR’s longitudinal structure and linkage potential also position it as a valuable resource for developing and externally validating prognostic models for oral disease outcomes such as tooth loss, an area where existing models have been limited by small sample sizes, inappropriate handling of missing data, and lack of external validation [47]. The utilization of the ODR represents a large-scale effort to integrate electronic oral health records within a major academic medical center in Singapore. This registry provides a valuable platform for research and healthcare service evaluation. Future expansion of its data scope and linkages will further enhance its utility, supporting quality assessment, care improvement, and targeted interventions aimed at reducing the burden of oral disease nationally.

## Data Availability

The data that support the findings of this study are available from National Dental Centre Singapore. Restrictions apply to the availability of these data, which were used under license for this study. Data are available from the authors upon reasonable request, with the permission of National Dental Centre Singapore.

## Acknowledgements

We also thank Bee Tin Goh, Chu Guan Koh, Li Qian, Dian Yi Chow, and Huihua Li for their valuable contributions and support in the development and implementation of the Oral Disease Registry.

## Availability of data

The data underlying this study cannot be shared publicly due to ethical and legal restrictions. The NDCS-ODR contains patient-level clinical records derived from electronic health records at the National Dental Centre Singapore (NDCS). Data access is governed by Singapore’s Personal Data Protection Act, the Human Biomedical Research Act, and SingHealth institutional guidelines on research data use. These restrictions are imposed by the NDCS Oral Disease Registry Executive Committee, which serves as the data access committee. Researchers interested in accessing ODR data must submit a proposal outlining their intended analysis, which is circulated to ODR investigators for feedback and potential collaboration. Upon endorsement, a formal data request is submitted to the ODR Executive Committee for approval.

## Captions

**S1 Table.** Detailed distribution of periodontal diagnoses in the ODR.

**S2 Table.** Number of periodontal visits by treatment type and year.

**S3 Table.** Completeness of periodontal charting at the site level by parameter.

**S4 Table.** Housing types, income ceiling for eligibility to purchase, and indicative pre-grant prices used to contextualise the Singapore Housing Index (SHI).

## Notes

### Competing Interest Statement

The authors have declared no competing interest.

### Funding Statement

Yes

### Author Declarations

The registry was established under SingHealth Centralised Institutional Review Board (CIRB) exemption 2022/2167 with a waiver of informed consent because only routinely collected, anonymized clinical data were used.

### Summary of Updates

This version has been updated to correct author name spellings and revise the author order. Minor edits have also been made to improve clarity and consistency throughout the manuscript.

